# A standardized system and App for continuous patient symptom logging in gastroduodenal disorders: design, implementation, and validation

**DOI:** 10.1101/2021.09.06.21263001

**Authors:** G Sebaratnam, N Karulkar, S Calder, JST Woodhead, C Keane, D Carson, C Varghese, P Du, S Waite, J Tack, CN Andrews, E Broadbent, A Gharibans, G O’Grady

**Author notes:** **Corresponding Author** Prof. Greg O’Grady, PhD, FRACS, Department of Surgery, University of Auckland, Auckland, New Zealand.

## Abstract

**Background:** Functional gastroduodenal disorders include functional dyspepsia, chronic nausea and vomiting syndromes, and gastroparesis. These disorders are common, but their overlapping symptomatology poses challenges to diagnosis, research, and therapy. This study aimed to introduce and validate a standardized patient symptom-logging system and App to aid in the accurate reporting of gastroduodenal symptoms for clinical and research applications.

**Methods:** The system was implemented in an iOS App including pictographic symptom illustrations, and two validation studies were conducted. To assess convergent and concurrent validity, a diverse cohort with chronic gastroduodenal symptoms undertook App-based symptom logging for 4-hours after a test meal. Individual and total post-prandial symptom scores were averaged and correlated against two previously validated instruments: PAGI-SYM (for convergent validity) and PAGI-QOL (for concurrent validity). To assess face and content validity, semi-structured qualitative interviews were conducted with patients.

**Key Results:** App-based symptom reporting demonstrated robust convergent validity with PAGI-SYM measures of nausea (*r*_S_=0.68), early satiation (*r*_S_=0.55), bloating (*r*_S_=0.48), heartburn (*r*_S_=0.47), upper gut pain (*r*_S_=0.40) and excessive fullness (*r*_S_=0.40); all *p<0*.*001* (n=79). The total App-reported Gastric Symptom Burden Score correlated positively with PAGI-SYM (*r*_S_=0.56; convergent validity; *p<0*.*001*), and negatively with PAGI-QOL (*r*_S_=-0.34; concurrent validity; *p=0*.*002*). Interviews demonstrated that the pictograms had adequate face and content validity.

**Conclusions and Inferences:** The continuous patient symptom-logging App demonstrated robust convergent, concurrent, face, and content validity when used within a 4-hour post-prandial test protocol. The App will enable standardized symptom reporting and is anticipated to provide utility in both research and clinical practice.

## Introduction

Functional dyspepsia (FD) and chronic nausea and vomiting syndromes (CNVS) affect 7.2% and 1.2% of the global population respectively, and significantly impact quality of life (1–3). According to the Rome IV Criteria, FD is characterized by excessive fullness and early satiation (dominant in the post-prandial distress syndrome subtype), and epigastric pain and/or burning (dominant in the epigastric pain syndrome subtype), while CNVS patients predominantly experience nausea and vomiting (4). However, these syndromes and symptoms often co-exist, while also overlapping with gastroparesis, which is controversially distinguished by the presence of delayed gastric emptying (5,6). Additional symptoms such as bloating and belching are commonly present in affected patients (4), while FD frequently also co-exists with gastro-oesophageal reflux disease (GERD) (7).

Distinguishing these disorders remains challenging owing to these overlaps and the ongoing lack of objective and specific biomarkers. A clear clinical characterization of specific symptoms is therefore essential for diagnosis, together with the exclusion of organic pathologies. A distinction must also be made between other potentially co-existing functional / gut-brain-axis disorders such as irritable bowel syndrome (8). Moreover, accurate characterisation is subject to the quality of clinical communication and may be negatively impacted by use of jargon, constraints on clinical time, and inaccuracy in patient recall of their symptom experiences (9). Pictograms have been shown to assist in the understanding and communication of gastric symptoms between patients and healthcare providers, improving symptom reporting accuracy (9,16). However further validation of pictogram use is desirable before they can be reliably integrated into clinical practice.

Validated instruments based on symptom recall are already available for longer-term assessments and are commonly used in research contexts, such as the Patient Assessment of Upper Gastrointestinal Symptom Severity Index (PAGI-SYM), Gastroparesis Cardinal Symptom Index (GCSI) and GCSI Daily Diary (10–12). However, a continuous reporting tool that enables construction of a real-time symptom profile is also desirable to allow comparison with concurrent diagnostic tests, investigate provocations, and evaluate interventions. Continuous granular symptom profiling is also particularly important in functional gastrointestinal (GI) disorders because temporal and severity correlations comprise part of the ‘Plausibility Criteria’ that are recommended for use in the evaluation of candidate pathophysiological mechanisms and emerging biomarkers (15).

The aim of this study was therefore to introduce and validate a patient symptom-logging system to aid patients and clinicians in the accurate reporting of gastroduodenal symptoms, including the use of pictograms. Once conceptualized, the standardized patient symptom-logging system was implemented in an iOS App and cloud-based reporting portal, before assessment of the convergent, concurrent, face and content validity in patient cohorts.

## Material and Methods

Ethics approval was granted by the Auckland Health Research Ethics Committee and the Conjoint Health Research Ethics Board at Calgary. All patients provided written informed consent. The study was reported per the Strengthening the Reporting of Observational studies in Epidemiology (STROBE) statement and Standards for Reporting Qualitative Research (SRQR) (17,18).

### Gastroduodenal Symptom Reporting System

A gastroduodenal symptom logging system was conceptualized as depicted in **Fig. 1**. Ten symptoms were selected for logging based on those covering the spectrum of functional gastroduodenal disorders (4,19); i.e. epigastric pain, epigastric burning, early satiation, excessive post-prandial fullness, nausea, vomiting, bloating, belching, heartburn, and reflux. Symptoms were divided into those that are continuously experienced vs discrete events (**Fig. 1A**). The design required patients to log symptoms at minimum 15-minute intervals, or more frequently if symptoms changed, including around a test meal. Intervals of 15 minutes have previously been established as sufficient to generate accurate gastroduodenal symptom profiling (14,20) (**Fig. 1B**). Discrete symptom events as well as continuous symptom data were reported graphically (**Fig. 1C**).

**Figure 1.**
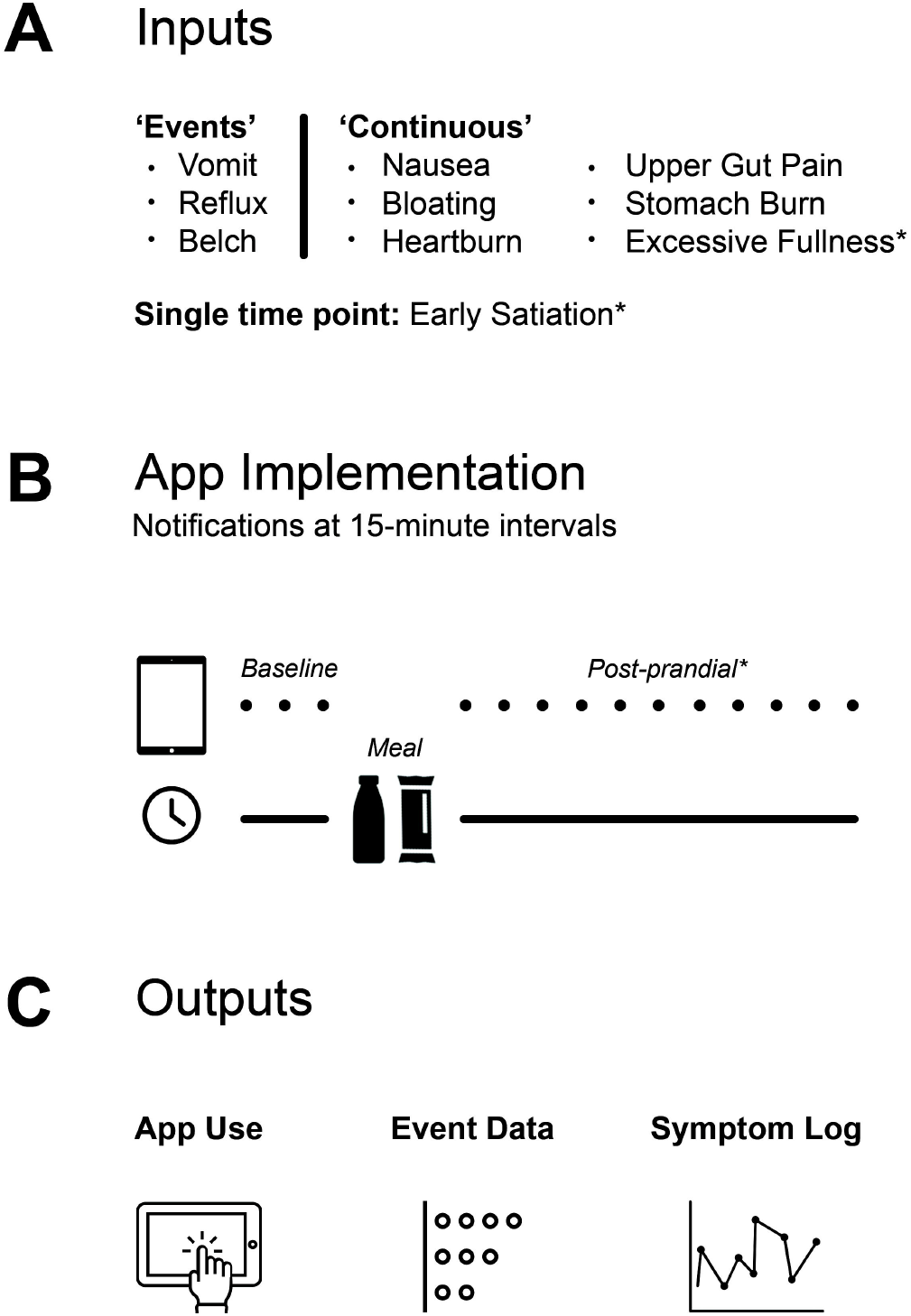
Pictorial depiction of the gastroduodenal symptom logging system. *Refers to symptoms logged only after a test meal.

### App Implementation

After the system was conceptualized, a custom App was implemented in iOS using the Swift 5 programming language, being designed to run on an iPad mini (Apple; Cupertino, California, USA). The App was developed by Alimetry (Auckland, New Zealand). The App allowed users to define fasted and/or fed testing durations and to specify a test meal if desired. Symptom reporting was standardized using both pictograms and written descriptors. A pictogram was assigned to each gastroduodenal symptom, being modified from those previously validated in a Belgian FD cohort by Tack et al (9) (**Fig. 2A and Supplementary Fig. 1**). The modifications were undertaken by designers with oversight from clinicians working in the field of GI motility to maintain content validity. The written descriptors were designed to be brief, jargon-free, and employed commonly accepted clinical terminology (**Fig. 2B**). Epigastric pain was simplified to ‘upper abdominal pain’, and epigastric burning to ‘stomach burn’.

**Figure 2.**
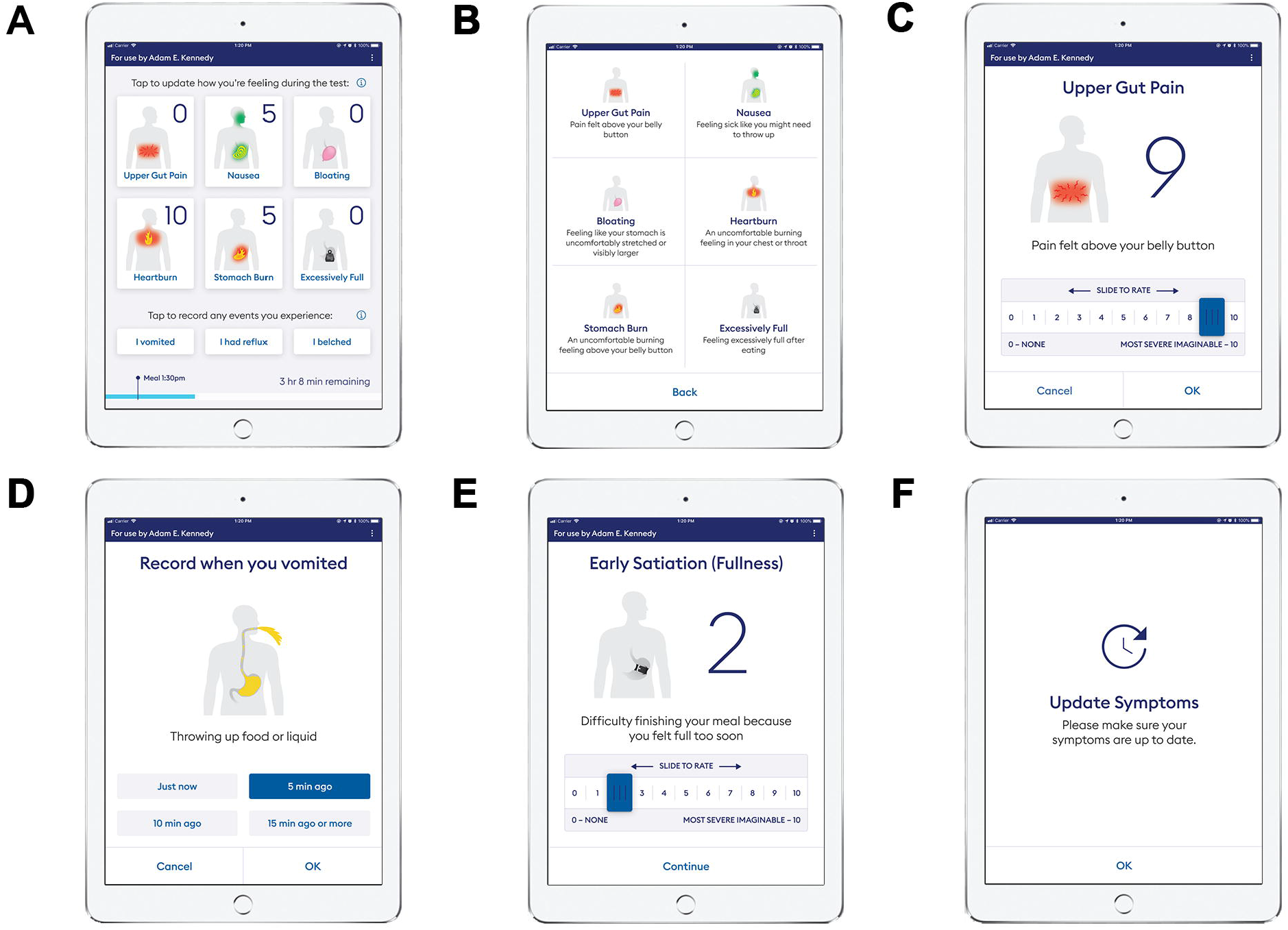
Screenshots of the iOS Symptom Logging App. **A)** Post-meal symptom dashboard display; **B)** Symptom explanations display; **C)** Upper gut gain symptom update display; D) Vomiting event logging display; **E)** Early satiation symptom update display; **F)** Symptom update reminder display. Content © Alimetry Ltd 2020, provided with permission.

The severity of each continuous symptom was assessed using a 0-10 Likert scale, with anchors at 0 ‘none’, indicating no symptom experience, and 10 indicating the ‘most severe imaginable’ extent of a symptom experience (**Fig. 2C**). This scale was chosen based on recommended guidance from the FDA (22), and because this scale is sensitive to clinically relevant changes in chronic pain intensity (21). Events were assessed using a two-step logging interface, allowing users to enter a type of event (**Fig. 2A)**, then the timing (**Fig. 2D**). Excessive fullness was only assessed postprandially, while early satiation was only assessed at a single time point immediately following the meal (**Fig. 2E**). The App displayed notifications every 15 minutes to alert the user to update their symptoms (**Fig. 2F**). Interactions with the App were also continuously tracked to ensure symptom logging completeness and compliance. At the end of the test, the data were automatically transferred from the iPad mini to a Health Insurance Portability and Accountability Act (HIPAA) compliant cloud server (Alimetry; Auckland, New Zealand), for automated generation of a graphical report in a secure online portal (**Fig. 3**).

**Figure 3.**
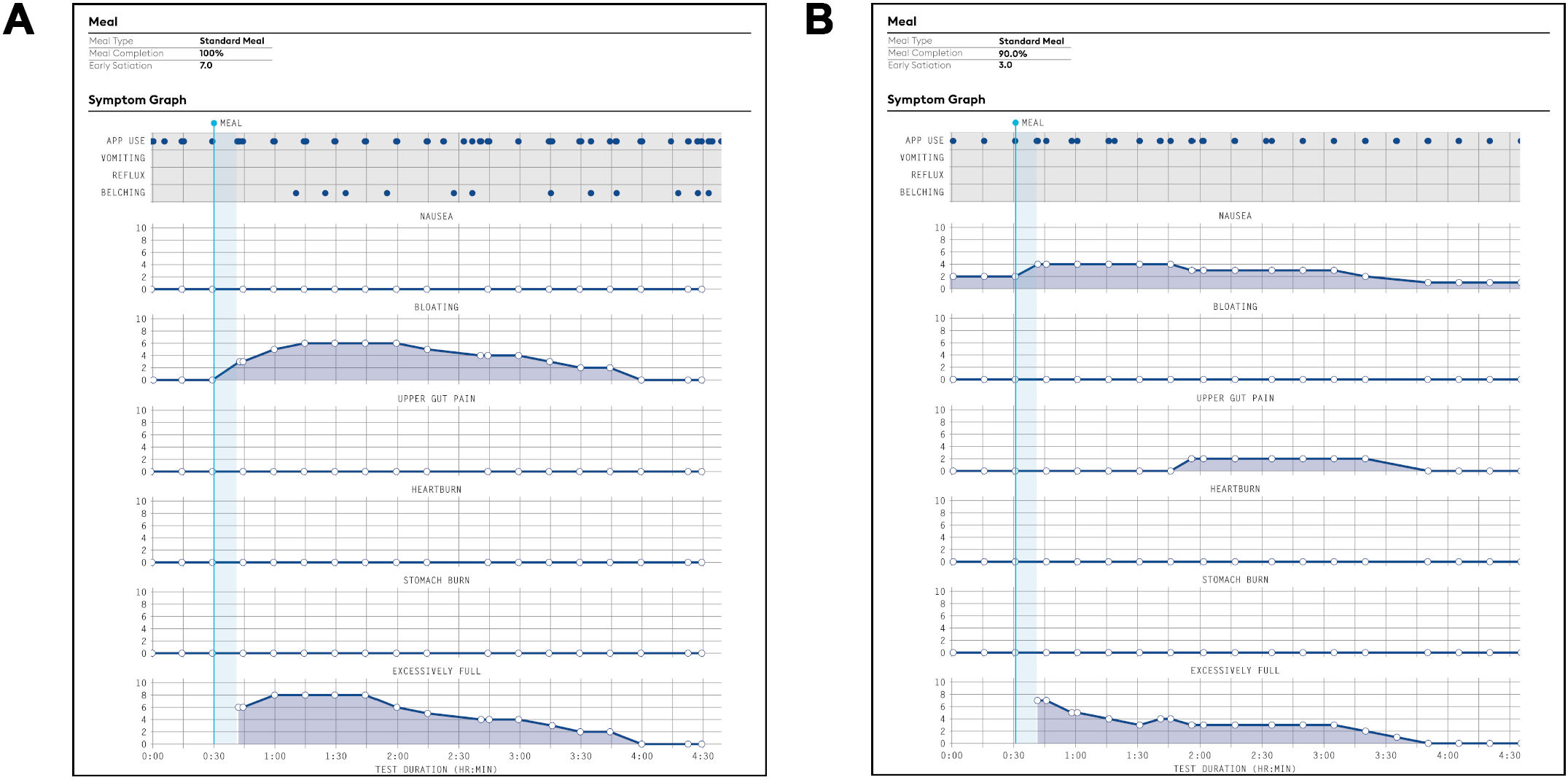
Examples of gastric test symptom reports following meal consumption in; **A)** A participant with functional dyspepsia participant; and **B)** A participant with chronic nausea and vomiting syndrome. Content © Alimetry Ltd 2020, provided with permission.

### Validation Study Design

Two validation studies were performed to assess the validity of the completed continuous symptom-logging App and its pictograms. Convergent and concurrent validity were evaluated in a multi-centre, observational cohort study of patients with chronic gastroduodenal symptoms by comparison with the validated PAGI-SYM and PAGI-QOL instruments. It was hypothesized that the App data would show positive correlations with the PAGI-SYM longer-term measure of gastric symptomatology (convergent validity), whereas for concurrent validity it was expected that the App data would be negatively correlated with the PAGI-QOL. Semi-structured patient interviews were used to assess face and content validity of the pictograms.

#### Study 1: Convergent and Concurrent Validation Study

Patients suffering chronic gastroduodenal symptoms were recruited from outpatient services or gastric scintigraphy referral lists. Patients recruited from outpatient services were referred with a diagnosis of gastroparesis, CNVS, or FD, while patients from scintigraphy referrals had chronic gastroduodenal symptoms without further differentiation, thereby ensuring the inclusion of a broad subset of eligible symptomatic patients typical of real-world clinical practice. Participants were excluded if they were aged <18 years, pregnant, or had an identified organic cause for their symptoms including metabolic or endocrine disorders, active GI infections, inflammatory bowel disease, or GI malignancy. All medications known to modify gastrointestinal motility were withheld for 48 hours prior to the study. A correlation coefficient of r > 0.3 was chosen as reasonably indicating validity, and a power calculation showed that a sample size of 79 participants would be needed to detect a difference in the App and the PAGI-SYM and PAGI-QoL outcomes with 80% power, a significance level of 0.05, and an effect size of r = 0.31.

The study protocol comprised a 30-minute fasted period, followed by consumption of a standardized meal over 10 minutes, followed by a 4-hour postprandial symptom-logging period. Patients referred from clinics received a nutrient drink (230 mL Ensure; Abbott Nutrition, IL, USA) and an oatmeal energy bar (250 kcal with 5 g fat, 45 g carbohydrate, 10 g protein, 7 g fibre; Clif Bar & Company, CA, USA), whereas patients recruited from scintigraphy lists received a standard egg meal (255 kcal with 72% carbohydrate, 24% protein, 2% fat, 2% fibre) or tofu equivalent if they had an egg allergy. All patients underwent a minimum six-hour pre-test fast.

The PAGI-SYM and PAGI-QOL were completed immediately prior to the start of the 30-minute fasted period. The PAGI-SYM is a 20-item validated questionnaire that asks participants to recall the severity of symptoms experienced over the last 2-weeks using a 6 point Likert scale from “none” to “very severe” (10). The PAGI-SYM consists of 6 subscales: i) heartburn / regurgitation; ii) postprandial fullness / early satiation; iii) bloating; iv) nausea / vomiting; v) lower abdominal pain, and vi) upper abdominal pain. Higher scores reflect higher symptom severity and burden (10). The PAGI-QoL is a disease-specific health-related quality of life patient-reported outcome measure (24). It consists of 30 items and 5 subscales: i) daily activities; ii) clothing; iii) diet and food habits; iv) relationships; v) psychological wellbeing and distress (24). It also asks participants to recall the previous 2 weeks’ experiences and uses a 6-point Likert scale. Throughout the study period participants used the App to log their symptoms as described above. At the end of the study participants also completed a single 5-point Likert scale to assess the ease of use of the App (0 = Very easy; 1 = somewhat easy; 2 = neutral; 3 = somewhat difficult; 4 = very difficult).

Statistical analyses were performed in GraphPad Prism v.9.1.2 (GraphPad; San Diego, California, USA). The PAGI-SYM summary score was calculated by taking the mean of all subscale scores (10). The PAGI-QOL summary score was calculated as the mean of all subscales, when all item responses were reverse coded (24), such that a higher PAGI-QOL score represents better disease-specific quality of life. Participant engagement with the App was assessed as the average time elapsed between App interactions, with non-compliance defined as median time exceeding 30 minutes between symptom logs. Before coding the App, a test was conducted on 5 patients with the symptom logging screens printed on paper to confirm that the system was working appropriately. Data from these patients were therefore excluded from the App usability and compliance testing data. Symptom logging data was taken from the 4-hr postprandial period to reflect when participants most likely experience symptoms in their everyday lives (25). The mean score and the area under the curve (AUC) were calculated for each symptom that was continuously logged during the 4-hour postprandial period, or as a single time-point for early satiation. A total symptom burden (referred to as the ‘Gastric Symptom Burden Score’) was also calculated, as both the sum of each participant’s mean symptom scores with early satiation and as the sum of individual symptom AUCs excluding early satiation. Spearman’s correlations were assessed between the Gastric Symptom Burden score obtained using each of these two methods. Discrete ‘events’ captured by the App (vomiting, reflux and belching) were not included in the individual symptom analysis nor the Gastric Symptom Burden score.

Spearman’s correlations were calculated to assess convergent validity between the post-prandial App-derived symptom severity scores (early satiation, bloating, upper gut pain, heartburn, excessive fullness and nausea) and the outcome of the corresponding PAGI-SYM item. Spearman’s correlation was also used to assess the association between the Gastric Symptom Burden score and the PAGI-SYM summary score (convergent validity) and gastric-specific quality of life as measured by the PAGI-QOL (concurrent validity). P<0.05 was considered statistically significant. Weak, moderate, and strong correlations were defined as r values greater than 0.1, 0.3 and 0.5 respectively.

#### Study 2: Face and Content Validation Study

Adults aged ≥ 18 years with a diagnosis of gastroparesis, CNVS, or FD (as per Rome IV), who resided in New Zealand and were able to provide informed consent, were eligible for the qualitative interview study. Potential participants were recruited via social media advertising, patient peer support groups, and clinical referrals. Exclusion criteria included the inability to speak or read English, and vulnerable participants (e.g. prisoners, individuals with a known cognitive impairment). Recruitment and data collection occurred between June 2020 and July 2021. We aimed to recruit between 5 - 15 participants in line with previous pictogram validation studies (9,26).

Semi-structured interviews were conducted using a web-based conferencing platform by two researchers trained and experienced in qualitative research methods. Participants were invited to have a support person present and interviews were scheduled according to participant request to ensure optimal conditions including privacy. During the interview, one pictogram was presented at a time and participants were asked to describe the symptom that was represented by the pictogram. After all pictograms had been presented, the exercise was repeated but participants were asked to choose the symptom label that they thought best represented the symptom depicted in the pictogram from a prescribed list.

Finally, there was a general discussion regarding any pictograms that were deemed to be poor representations, any symptoms that had not been well represented, and any recommendations for improvement. The interviews were recorded and transcribed by one of the interviewers with participant consent. Missing responses were excluded from analysis.

An adapted form of iterative thematic analysis, with a semi-quantitative approach, was utilised to evaluate participant responses. Agreement between the participants’ interpretation of the symptom pictogram and the ‘intended symptom’ was calculated and is reported as a percentage. Participant commentary was coded for each pictogram and analysed for common themes. Two coders, including one who did not participate in the interviews, coded all participant responses based on *a priori* coding categories. Coding agreement was assessed after transcription of the first two interviews and after coding of all interviews.

## Results

### Study 1: Convergent and Concurrent Validity

A total of 79 patients were included with median age 41 years (*IQR* = 27-52), most being female (82%), and a majority self-identifying as Caucasian (71%). All tests except one were performed in the morning. Two subjects who completed the face and content validity study below were subsequently also recruited into the convergent and concurrent validity study. **Table 1** provides a detailed description of participant characteristics. Patients had a representative mix of FD (12.7%), CNVS or gastroparesis (53.2%), and chronic gastroduodenal symptoms not further delineated (scintigraphy referral group; 34.2%). Across the whole cohort, 16.5% had a concurrent diagnosis of irritable bowel syndrome, and 26.6% had a concurrent psychological comorbidity (anxiety, depression, and/or post-traumatic stress syndrome).

On average, participants interacted with the App at a median of 8.5-minute intervals (*IQR =* 6.5-10.2), indicating high engagement. The average time between symptom logs did not exceed 13.6 minutes in any patient, demonstrating a 100% compliance rate with symptom logging. Of all participants who used the app, 90.1% reported that it was ‘very easy to use’ and 9.9% reported it was ‘somewhat easy’. Three participants did not respond to the ease-of-use question.

Example symptom report outputs are provided in **Fig. 3A &B** for patients with FD and CNVS respectively. Across all subjects, strong correlations were found between the mean and AUC metrics for the App post-prandial symptom scores, as well as for the calculated Gastric Symptom Burden Score (*r*_s_ = 0.95 - 0.99). The validity comparisons therefore only employed the mean scores, with the AUC metrics not being further employed.

Moderate to strong associations were found for all assessed symptoms (*p* < 0.001) between the App and PAGI-SYM individual symptom measures (**Fig. 4**). Early satiation (*r*_s_ = 0.55) and nausea (*r*_s_ = 0.67) correlated strongly, while upper gut pain (*r*_s_ = 0.40), heartburn (*r*_s_ = 0.47), bloating (*r*_s_ = 0.42), and excessive fullness (*r*_s_ = 0.40) demonstrated moderate associations with their PAGI-SYM equivalent (**Fig. 4**). There was a strong association between the total Gastric Symptom Burden Score and the mean PAGI-SYM summary score (*r*_s_ = 0.56, *p* < 0.001) (**Fig. 5A**). These results confirm convergent validity between the App and the PAGI-SYM questionnaire.

**Figure 4.**
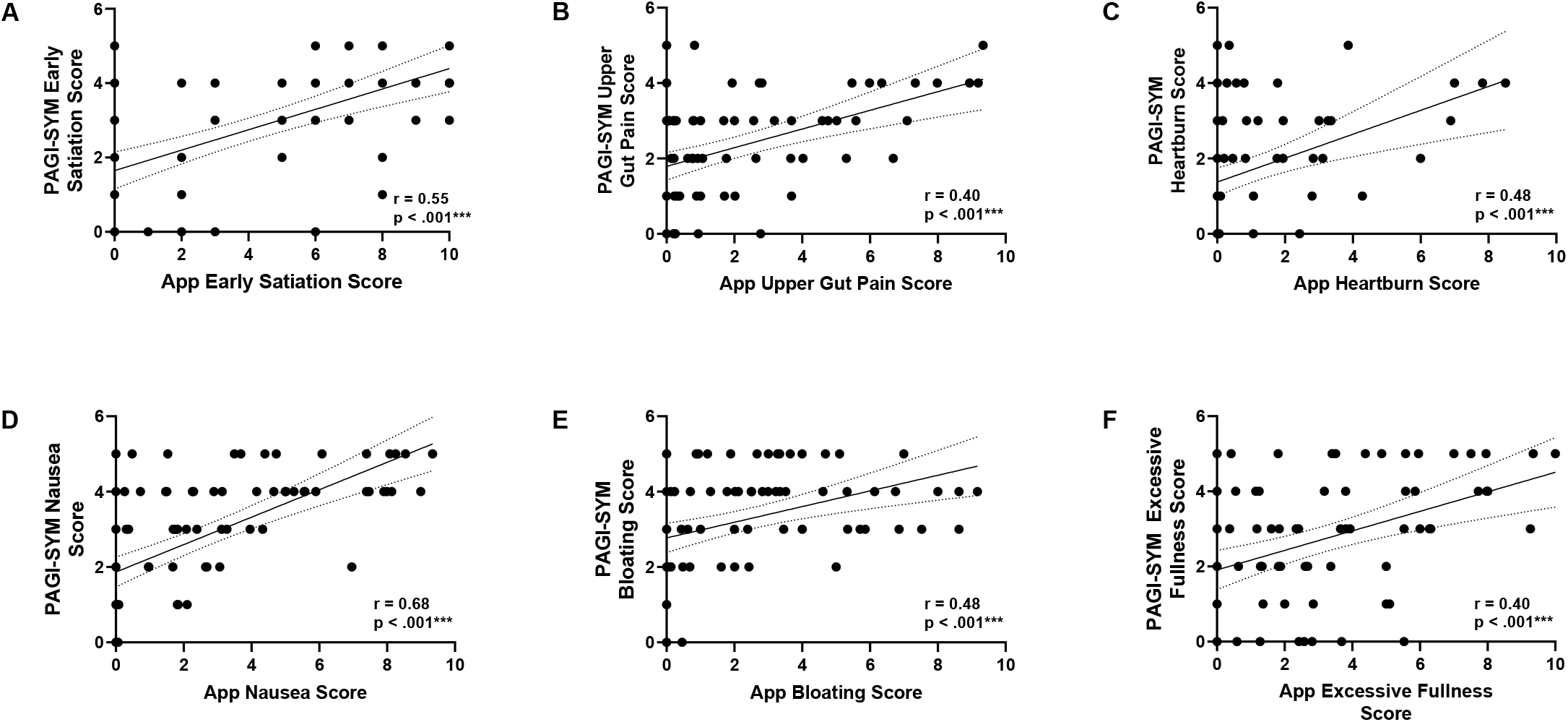
Scatterplots showing correlations between App symptom scores and related PAGI-SYM item scores (n=79). Dotted lines represent the corresponding 95% confidence intervals.

**Figure 5.**
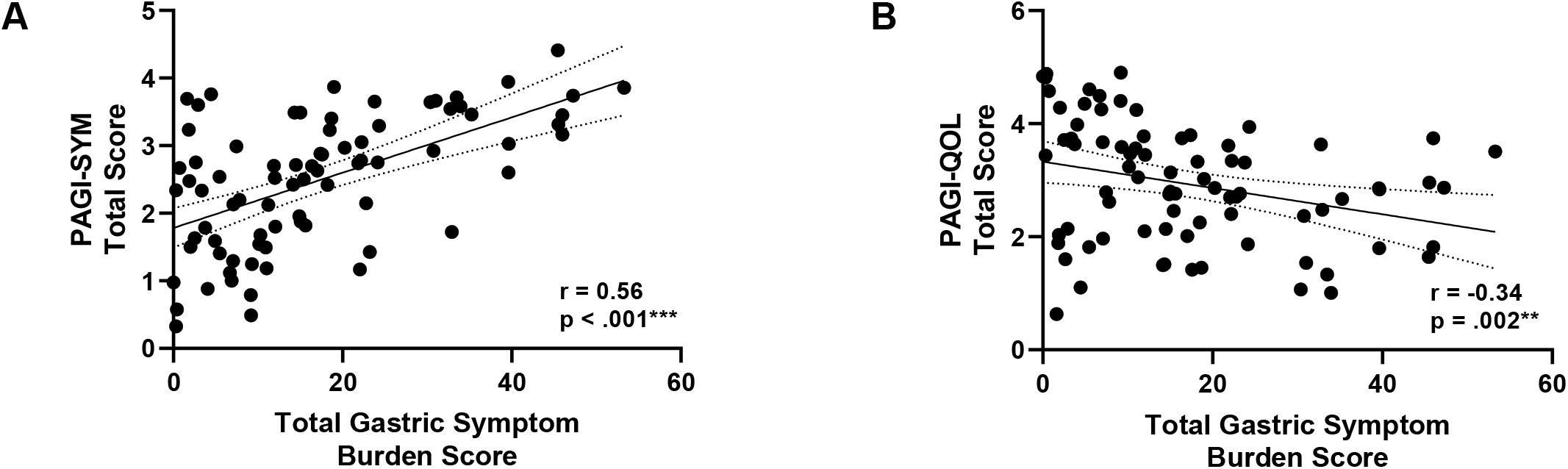
Scatterplots showing correlations between the total Gastric Symptom Burden Score reported via the App vs **A)** PAGI-SYM overall score; and **B)** PAGI-QOL overall score (n=79). Dotted lines represent the corresponding 95% confidence intervals.

The Gastric Symptom Burden Score had a significant, moderate, negative association with the PAGI-QOL summary score (*r*_s_ = -0.34 *p* = 0.002) (**Fig. 5B**). This finding confirms concurrent validity, as participants who reported a higher burden of postprandial gastric symptoms in the App were also likely to report reduced quality of life.

### Study 2: Face and Content Validity

Eight patients participated in the pictogram validation interviews. However, one interview was excluded due to loss of data, and a second was excluded as the participant had completed the convergent and concurrent validity study prior to their interview and were therefore not pictogram naive. The analysed interview cohort consisted of Caucasian females of median age 38 years (IQR = 25.5 – 46.0), all with CNVS or gastroparesis. The interview length ranged from 9 to 36 minutes.

The highest performing pictogram was that depicting vomiting, which had 100% agreement, meaning all participants suggested that the pictogram described vomiting as it was intended to do. Pictograms for belching (83% agreement), upper abdominal pain (83%), heartburn (67%), bloating (67%), nausea (67%) and reflux (67%) showed high to moderate degrees of agreement. Early satiation (0%) and excessive fullness (16%) had poor participant agreement.

When participants were provided with the list of symptoms to match with the pictogram, agreement improved. Belching, heartburn, nausea, reflux, upper abdominal pain, and vomiting showed 100% agreement. Feeling excessively full (80% agreement), bloating (80%), and early satiation (50%) had strong to moderate agreement.

Assessment of patient verbal assessments of the pictograms contributed little further data beyond the above analyses, generally matching the summarised findings. The vomiting pictogram received the most positive feedback, whereas early satiation received the most negative feedback with four out of six participants stating that the pictogram did not match well with the symptom label. Suggestions regarding other pictograms (feeling excessively full, heartburn, nausea, and reflux) were heterogenous (example responses provided in **Supplementary Table 1)**.

## Discussion

This study reports the design, implementation, and validation of a standardized system, App, and cloud-based reporting portal for continuous patient symptom logging in functional gastroduodenal disorders. The App, coded in iOS, differentiates continuous symptoms from discrete symptom events and incorporates standardized symptom identifiers and scales for severity gradings. Robust correlations were shown between the App and PAGI-SYM measures of individual and overall gastric symptoms indicating convergent validity. The association between PAGI-QOL scores and App-reported symptoms reflected an expected relationship between gastric-specific quality of life and overall gastric symptom burden, demonstrating concurrent validity. Most patients reported the App was “very easy to use”, validating usability. Qualitative interviews demonstrated adequate face and content validity of the pictogram-based approach to symptom reporting, although finding some areas for improvement. Overall, these findings show that the App-based 4-hour ‘symptom snapshot’ following a standard meal is a valid, patient-centric, and representative method for evaluating gastroduodenal symptoms.

Previous literature has identified pictograms as an efficient way of communicating subjective symptoms, being superior to text alone (9,26,27). Tack et al. previously introduced a series of pictograms specific for functional GI disorders and demonstrated improved patient symptom reporting accuracy and symptom understanding in a cohort of FD patients (9). In the present study, these pictograms were modified and incorporated into our App to support standardized real-time symptom capture around a meal. Our qualitative data showed that the depiction of vomiting performed ideally, and those for belching, heartburn, upper abdominal pain, and bloating also performed strongly. However, further improvements are desirable to better depict early satiation and excessive fullness in subsequent iterations of the App.

A recent study by Kuwelker et al. similarly reported that symptoms captured throughout a 4-hour gastric scintigraphy study correlated well with validated measures of gastric symptoms in patients with diabetes mellitus (14). As such, the findings from the current study corroborate the validity of the 4-hour gastric symptom capture window and extend these findings to a greater range of patients with gastroduodenal symptoms (14). Together, these two studies provide a strong foundation for the external validity of utilizing a 4-hour gastric symptom snapshot to assess gastric symptomatology in clinical practice and research (14).

The PAGI-SYM questionnaire is one of the few generic functional gastroduodenal symptom severity metrics which has good test-retest consistency, is well-validated, and is easily administered (10). The GCSI Daily Dairy, employing a key subset of the PAGI-SYM items, was subsequently introduced as a validated method of daily symptom reporting (12). A small number of other tools exist, but their uptake is disease specific or not widespread (11,28,29). There is a paucity of standardized and validated techniques to enable the assessment of gastric symptoms continuously over a defined test period. This App will therefore fill a significant gap, offering broad utility. In clinical settings, it offers standardized accurate symptom reporting to aid patient-clinician communication, being particularly useful when symptom correlations are required for diagnostic tests involving provocations, such as nutrient drink tests of accommodation, gastric emptying tests, and body surface gastric mapping (14,30,31). The App is also anticipated to be useful in defining and comparing symptom experiences before and after interventions, and for pathophysiological studies employing the ‘Plausibility Criteria’ for mechanisms of functional GI disorders (15,20).

Some limitations to this study are acknowledged. While a relatively large multi-centre, international, prospective cohort of symptomatic patients were included, specific patient subgroups entering the study via scintigraphy referral lists were not completely defined by Rome Criteria or other diagnoses. This allowed demonstration of validity over a diverse patient cohort but precluded the assessment of criterion validity. Symptom correlations with PAGI-SYM were only possible for the continuous symptoms, and not the discrete symptom events. Two test meals were included, enabling validity to be demonstrated across a range of meal types, however, this may have diversified symptom profiling during the postprandial window. Additionally, within our cohort, all but one patient was studied in the morning, whereas recent data suggests that gastric symptom severity may be at its worst in the evening in gastroparesis patients (32). It is notable that despite these factors, symptom reporting via the App still demonstrated robust validity in capturing a reasonably typical symptom burden experience.

A limitation to the qualitative study was that the pictogram validation cohort consisted only of Caucasian women with CNVS or gastroparesis. Such patients are typical of CNVS and gastroparesis demographics in our practice, however, culture, gender, and disorder experiences could all possibly influence symptom interpretations (33–36). Self-reporting, recall, and social desirability biases are also possible due to the inherent nature of interview data (37,38). Nevertheless, the interviews revealed consistent and helpful insights including areas for improvement. The gastroduodenal symptom logging App will be iteratively improved over time, presenting future opportunities for re-evaluation in an increasingly diverse interview group.

In summary, this study introduces a continuous patient symptom logging system, App, and reporting tool for gastroduodenal disorders, and demonstrates that a 4-hour post-prandial symptom snapshot is a valid and representative approach to measuring symptom experiences and their burden. The system and App are anticipated to be a useful addition to clinical practice and research.

## Data Availability

Not available

## Acknowledgements and Funding

The authors would like to thank the patients who took part in this study and our clinical recruitment staff Gen Johnston, India Wallace, Lynn Wilsack and Renata Rehak. We thank Jim Hannon-Tan and Tom Russell for their invaluable input to the App and pictogram design. The validation studies were supported by the Health Research Council of New Zealand and the John Mitchell Crouch Fellowship from the Royal Australasian College of Surgeons.

---

**Supplementary Figure 1.**
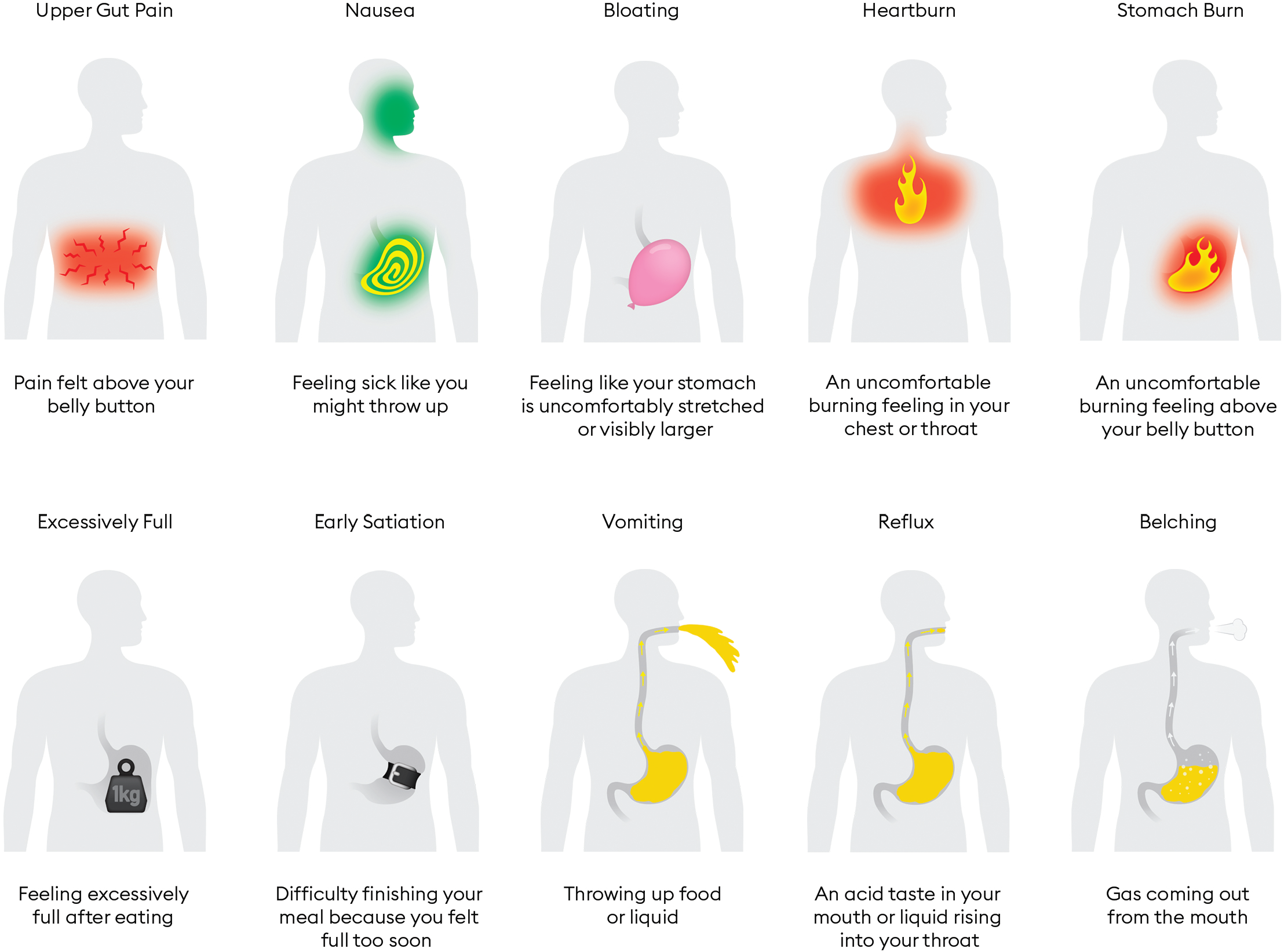
Illustrations of all Pictograms and descriptions used in this study. Content © Alimetry Ltd 2020, provided with permission.

