## Supplementary material for "A standardized system and App for continuous patient symptom logging in gastroduodenal disorders: design, implementation, and validation": Table 1

Table 1: Participant Clinical and Demographic Characteristics.

| **Characteristic** | **FD (n=10)** | **CNVS or Gastroparesis (n = 42)** | **Scintigraphy (n = 27)** | **Total Sample (N=79)** |
|---|---|---|---|---|
| **Age** (years; *median)* (IQR) | *51* (32-64) | *34* (26-44) | *45* (31-56) | *41* (27-52) |
| **Female, n (%)** | 9 (90) | 37 (88) | 19 (70) | 65 (82) |
| **Ethnicity, n (%)** | | | | |
| Caucasian | 9 (90) | 31 (74) | 16 (59) | 56 (71) |
| Māori | 0 (0) | 3 (7) | 4 (15) | 7 (9) |
| Chinese | 1 (10) | 1 (2) | 1 (4) | 3 (4) |
| Pasifika | 0 (0) | 1 (2) | 1 (4) | 2 (3) |
| Indian | 0 (0) | 1 (2) | 1 (4) | 2 (3) |
| Other | 0 (0) | 5 (12) | 4 (15) | 9 (11) |
| **BMI** ($kgm^2$) *mean* (Std)[a] | *22.6* (4.8) | *24.1* (4.4) | 24.0 (4.5) | 23.9 (4.5) |
| **Comorbidities, n (%)** | | | | |
| Diabetes | 1(10) | 5 (12) | 1 (4) | 7 (9) |
| Hypo/Hyper-thyroidism | 1(10) | 0 (0) | 1 (4) | 2 (3) |
| Cardio-Respiratory | 1(10) | 8 (19) | 6 (22) | 15 (19) |
| IBS | 1(10) | 7 (17) | 5 (19) | 13 (16) |
| Anxiety / Depression / PTSD | 2 (20) | 16 (38) | 3 (11) | 21 (27) |

Table 1 continued

Table 1 Continued

| **Characteristic** | **FD (only) (n=10)** | **CNVS or Gastroparesis (n = 42)** | **Scintigraphy (n = 27)** | **Total Sample (n=79)** |
|---|---|---|---|---|
| **PAGI-SYM Score** *Mdn* (IQR) | *1.23* (1.03 - 1.73) | *2.45* (1.83 – 3.30) | *2.60* (1.40 - 3.45) | *2.40* (1.50 - 3.30) |
| **PAGI-QOL Score** *Mdn* (IQR) | *3.34* (2.74 - 4.39) | *2.68* (1.82- 3.44) | *3.50* (2.40 - 3.80) | *2.86* (2.03 - 3.71) |
| **GCSI Score** *Mdn* (IQR) | *1.71* (1.31 - 2.47) | *3.17* (2.52 – 3.67) | *2.83* (2.08 - 3.64) | *2.83* (2.06-3.64) |

Note: BMI = Body Mass Index; IBS = Irritable Bowel Syndrome; PTSD = post-traumatic stress disorder; GCSI = Gastroparesis Cardinal Symptom Index.
