## Supplemental Table 1 for "A standardized system and App for continuous patient symptom logging in gastroduodenal disorders: design, implementation, and validation"

Supplementary Table 1

Examples of patient responses to the symptom pictograms and recommendations for improvement.

| Intended Symptom | Examples of Participant Feedback to Pictogram |
|---|---|
| Belching | “...belching kind of thing”<br><br>"It's a difficult one to actually image without changing the person's shape" |
| Excessive Fullness | “stomach heaviness”<br><br>"maybe the feeling full, the stomach full and, maybe, you know, arrows pushing out to be like, you know, it's full" |
| Heartburn | “heartburn and acid going up through the back of your throat”<br><br>“Bad fit” |
| Nausea | “... that ‘yuck’ flushing feeling that you get when you’re feeling nauseous”<br><br>“I’d probably do a squiggly line as opposed to a circle-y line, that is probably more like hypnosis” |
| Reflux | “Vomiting slash maybe kind of reflux-y again”<br><br>"Maybe change the colour? Of the stomach?" |
| Upper Abdominal Pain | “I suppose that is abdo[minal] pain or yeah, a bit of a burning sensation in your stomach maybe” |
| Vomiting | “Definitely vomiting”<br>“vomiting. That is kind of obvious” |
| Bloating | “There’s a balloon, so it’s obviously bloating”<br>“... [the balloon should be] inside [the stomach]. I think that would be better yeah” |

(Supplementary Table 1 continued)

(Supplementary Table 1 continued)

| Intended Symptom | Examples of Participant Feedback to Pictogram |
|---|---|
| Early Satiety | “Restricted or tight stomach”<br><br>“The picture is a little harder than the other ones.... wording is a little harder than the other ones...and the combination of the two might miss some people”<br><br>“I don’t know how you would make a picture better than that one…maybe changing the wording might help people understand the picture better?” |
